# Behavioral profiles associated with adherence to adjuvant endocrine therapy in breast cancer: a retrospective population-based cohort study

**DOI:** 10.64898/2026.05.25.26353903

**Authors:** Aaron Dibner-Dunlap, Staci Sutermaster, Peter Smittenaar, Sema Sgaier

## Abstract

**Purpose:** Adjuvant endocrine therapy (AET) substantially reduces recurrence and mortality in hormone receptor-positive breast cancer but requires sustained daily adherence over 5 to 10 years. Approximately one-third of patients fall short of recommended adherence in the first year alone, largely due to distinct combinations of attitudes, barriers, and circumstances. Existing studies have catalogued individual risk factors but lack the scale and breadth to characterize how these factors co-occur within patients, or to distinguish behavioral drivers from confounding by clinical and demographic context. We sought to characterize the behavioral and social heterogeneity underlying AET adherence in a national real-world cohort. Moving beyond population-average risk factors, we identify the distinct patient profiles, and the differing drivers within them, that any effective adherence strategy must address.

**Methods:** We conducted a retrospective cohort study of US women with invasive breast cancer diagnosed between 2016 and 2025, linking two large-scale, individual-level datasets through privacy-preserving tokenization: Surgo Health’s BehavioralPulse™, which provides modeled individual-level behavioral and attitudinal risk scores together with consumer sociodemographic attributes, and longitudinal medical and pharmacy claims from a claims data provider. Eligible patients underwent 1 to 2 breast surgeries, initiated oral AET (tamoxifen or aromatase inhibitors), and maintained continuous insurance coverage for 365 days following therapy initiation. The primary outcome was adherence, defined as medication possession ratio (MPR) ≥80% in the first year. Mixed-effects logistic regression with a random intercept for ZIP3 estimated adjusted associations across behavioral, sociodemographic, and clinical predictors. To characterize how behavioral factors co-occur within patients, we identified the most prevalent configurations of the statistically significant behavioral predictors and estimated their relative association with adherence, holding clinical and demographic factors constant.

**Results:** The final cohort included 401,450 women, of whom 280,595 (69.9%) achieved MPR ≥80%. Several behavioral factors were independently associated with adherence after adjustment for clinical and demographic covariates, including comfort following medication instructions (aOR, 1.15; 95% CI, 1.06-1.24), geographic proximity to breast oncologists (aOR, 1.17; 95% CI, 1.04-1.32), tangible instrumental social support (aOR, 1.06; 95% CI, 1.00-1.13), religiosity (aOR, 1.04; 95% CI, 1.01-1.08), concern about sexual side effects (aOR, 0.96; 95% CI, 0.93-0.99), and cost-related access barriers (aOR, 0.97; 95% CI, 0.95-1.00). The 10 most common configurations of significant behavioral predictors accounted for over 70% of the cohort, with the two most prevalent representing more than 40% of patients. The most common profile, defined by an absence of behavioral barriers and the presence of social support, was associated with a positive behavioral contribution to adherence propensity (behavioral linear predictor OR = 1.18; 95% CI: 1.04-1.36) comparable in magnitude to several established clinical predictors. Compared against this referent profile, six of the nine remaining profiles had lower adherence, with relative odds ranging from approximately 0.92 (95% CI: 0.89-0.95) to 0.97 (95% CI: 0.94-0.99). One profile, similar to the reference but including high trust in doctors, was associated with higher adherence odds (1.04, 95% CI: 1.01-1.07). These profiles arose from substantively different underlying combinations of factors: segments dominated by cost barriers, by side-effect concerns, or by limited social support produced comparable overall adherence risk but through distinct pathways.

**Conclusion:** In this national cohort, nearly one-third of women did not achieve recommended first-year adherence to AET. The pathways to non-adherence were heterogeneous, structured into recurring behavioral profiles rather than randomly distributed across patients. This heterogeneity is clinically meaningful: patients with similar adherence risk may benefit from substantially different forms of support, ranging from financial navigation to side-effect management to social support resources. Surfacing this structure required linking individual-level behavioral data to large-scale claims data, offering a practical foundation for optimal design of patient-centered adherence interventions that are tailored to the specific configurations of barriers patients actually face.

## Introduction

The breast cancer patient journey is defined by a series of demanding milestones that are shaped not only by clinical characteristics, but also by the patient’s underlying behavioral and social factors. After diagnosis and the completion of active treatment, patients enter a period that requires high levels of personal agency to maintain long-term medication adherence and routine follow-up care [1]. A key component of long-term care for patients with hormone receptor-positive (HR+) breast cancer is adjuvant endocrine therapy (AET), which reduces the risk of recurrence. An estimated 200,000 patients initiate such treatment annually in the US [2–4].

Despite being highly effective at improving long-term survival [5], roughly half of patients discontinue AET prematurely or fail to take the medication as prescribed [6, 7]. Adherence is widely recognized as a priority across the healthcare system and has been the focus of intervention efforts from payers, providers, life science companies, and patient advocacy groups. Yet interventions designed to improve adherence have shown mixed results [8] and most critically, there is no evidence that adherence is improving over time [9]. Despite active drug development pipelines for adjuvant therapy [10], their population-level survival benefit is constrained by the same adherence problem that affects current therapies.

One approach to improve adherence is to better address the barriers faced by each patient individually. Patients that initiate AET may be similar in terms of their clinical characteristics, but they are highly diverse when it comes to their odds of successful adherence and the underlying reasons why. Behavioral factors are known to affect AET adherence. For example, self-efficacy and depressive symptoms are consistently listed as critical factors that impact AET adherence [11, 12]. Other risk factors are positive decisional balance (the belief that the treatment is necessary and outweighs the costs); the quality of relationship with providers; social support; concern over side effects; and forgetfulness [11, 12].

These factors are well-known individually, but characterizing how they combine within patients has been challenging due to methodological constraints: single-region cohorts that mask broader disparities across the US; geographically-aggregated estimates of behavioral drivers that suffer from ecological bias; and patient survey designs that rely on collecting additional data during an already demanding journey [4, 13–18]. To address these shortcomings, we developed BehavioralPulse™, a high-resolution dataset of modeled individual-level health attitudes, beliefs, and barriers for US adults. Linking these behavioral propensities with longitudinal claims enables patient-level analysis of a broad set of factors without adding patient burden.

In this study, we use large-scale, linked BehavioralPulse-claims data to test two related hypotheses. First, that behavioral factors are independently associated with adherence after adjustment for clinical and sociodemographic characteristics, such that two patients with otherwise identical clinical profiles may face meaningfully different odds of adherence. Second, that these drivers exist in recurring configurations, such that we can identify segments of patients with similar overall adherence risk that arrived there through substantively different pathways, each implying a different intervention need.

## Methods

### Data Sources and Linkage

We constructed a retrospective cohort by linking two large-scale, individual-level data sources using a privacy-preserving, hash-based tokenization approach [19, 20]. The first source was BehavioralPulse™, a national-scale dataset of behavioral propensity scores. It estimates, for the majority of US adults, a wide range of health beliefs, perceived barriers, social support, trust in clinicians, and related constructs (see Appendix for methodology behind BehavioralPulse). This dataset also included appended consumer-level sociodemographic attributes, including estimated household income, educational attainment, homeownership, marital status, occupation, and household composition.

The second source consisted of large-scale public and commercial claims data from the Komodo Healthcare Map® for US women with at least one breast cancer diagnosis recorded between January 2016 and April 2025. Claims included inpatient, outpatient, and pharmacy records, enabling identification of surgical procedures, endocrine therapy prescriptions or injections, comorbid diagnoses, and insurance coverage continuity. Prior to applying cohort eligibility criteria, the linked dataset comprised approximately 2.1 million unique women.

### Cohort Definition

Analyses focused on female patients who initiated standard oral AET following surgical treatment for invasive breast cancer, and who had sufficient observable followup to assess one year of adherence behavior. Women were required to have at least two claims containing an ICD-10 diagnosis code for invasive breast cancer (C50.*) at any point in their claims history to reduce the likelihood of miscoding. Eligible patients were further required to have undergone one or two breast cancer-related surgeries (mastectomy or lumpectomy). For patients with two surgeries, both were required to occur within one year of each other, reflecting common clinical scenarios such as re-excision or conversion to mastectomy after an initial lumpectomy; in these cases, the latter surgery was treated as the index surgery. We excluded patients with three or more surgeries, or those with two surgeries more than 365 days apart (6.3% of surgical patients) as those were more likely to reflect medically complex histories with potentially complicated treatment trajectories.

Patients were required to have evidence of AET following surgery, defined by an oral endocrine therapy prescription (tamoxifen or aromatase inhibitors [AIs]). The date of the first observed postsurgical AET prescription or administration served as the index date for adherence measurement; the index date was required to fall between March 1 2016, to establish minimal comorbidity observability window, and April 1, 2024, to establish the minimum adherence observability window. Patients could have received neoadjuvant endocrine therapy prior to surgery, which was retained as a covariate.

To ensure complete observation of adherence behavior and to partially mitigate bias from unobserved mortality, patients were required to have continuous insurance coverage during the 365 days following AET initiation. We excluded patients with evidence of ovarian function suppression (e.g., goserelin or leuprolide; 6.9% of patients with observed AET), given the distinct treatment paradigms and adherence considerations in premenopausal disease. As age of menopause onset varies, and menopause status is not reliably reported in claims, we applied no other restrictions regarding menopause status. The final analytic cohort comprised 401,450 women (Table 1).

**Table 1:** Inclusion criteria and participant flowchart. AET: adjuvant endocrine therapy.

| Criteria | Sample Patients | Excluded from Prior Step |
| --- | --- | --- |
| Initial analytic dataset | 2,169,159 | -- |
| 2+ breast cancer diagnoses | 1,867,518 | 301,641 |
| 1-2 surgeries within 365 days | 724,888 | 1,142,630 |
| Oral AET initiated | 447,446 | 277,442 |
| Continuous Insurance Coverage | 415,552 | 31,894 |
| AET Index date prior to 1 April 2024 | 402,088 | 13,464 |
| Removal of <1% missing observations | 401,450 | 638 |

### Outcome Definition

The primary outcome was adherence to AET, defined as achieving a medication possession ratio (MPR) of at least 80% during the 365 days following initiation of postsurgical endocrine therapy. MPR was calculated using pharmacy dispensing dates and days’ supply, allowing construction of a longitudinal supply timeline and identification of delayed or missed refills. Adherence was operationalized as a binary outcome (≥80% vs <80%), consistent with prior studies focusing on AET adherence [21–25].

### Predictors

We selected predictors from claims, consumer sociodemographic data, and behavioral risk measures. We selected behavioral predictors from the BehavioralPulse dataset using a two-stage process. First, we identified variables with strong conceptual or empirical relevance to medication adherence based on prior literature, such as concerns about side effects, cost-related access barriers, social support, religiosity, and health literacy. Second, to mitigate multicollinearity, we iteratively removed variables with pairwise Pearson correlations exceeding 0.7, using a prespecified prioritization ranking drawing from conceptual relevance, until all remaining correlations were below 0.7. Fifty behavioral variables remained after this process. We binarized behavioral risk scores at the population mean to facilitate interpretability, reflecting the presence or absence of a given belief or barrier.

Claims-derived variables included age, race and ethnicity, insurance type (commercial vs Medicare), state and 3-digit ZIP of residence, and comorbid conditions documented prior to AET initiation, including anxiety, depression, arthralgia, osteoporosis, vasomotor symptoms, and sexual dysfunction. We also included an indicator for prior endocrine therapy exposure (tamoxifen or AIs). As data were derived from administrative claims, we were not able to ascertain other clinical confounders such as tumor stage, nodal status, or other nuances in treatment regimen commonly captured in clinical records but excluded from claims [26].

Several predictors exhibited minimal missingness (<1%), such as patient state, 3-digit ZIP, or age; we excluded observations missing these variables. For variables with moderate missingness, particularly sensitive sociodemographic attributes such as race and ethnicity (13% missing), we imputed missing values to zero (for numeric predictors) and included an explicit missingness category for categorical predictors. We chose this approach over casewise deletion because the missing data are unlikely to be completely random (non-MCAR); deletion would have introduced potentially significant bias. A sensitivity analysis evaluated the robustness of this approach.

### Statistical Analysis

We first set out to identify significant behavioral predictors, which were then used to define the most common behavioral profiles. We estimated a mixed-effects logistic regression model with a random intercept for ZIP3 to account for geographic clustering. We estimated adjusted odds ratios (aORs) with 95% confidence intervals (CIs) to identify behavioral predictors with a significant association with adherence, after adjusting for all other demographic and clinical predictors. We then took this subset of significant behavioral factors, and identified their 10 most common patient profiles present across the patient population. For each profile, we fixed the remaining behavioral predictors at their conditional mean and held all other demographic and clinical predictors constant. We estimated how likely each patient profile was to adhere to AET by drawing coefficients from a multivariate normal distribution defined by the estimated coefficient vector and covariance matrix [27]. Using the most common profile as reference, we used Monte Carlo simulation to obtain the ORs and CI for adherence by the other 9 profiles [28].

We assessed model discrimination using the area under the receiver operating characteristic curve (ROC-AUC) in a 30% held-out test set. Explained variance was summarized using Nakagawa’s marginal and conditional R^2^, and intraclass correlation coefficients quantified the contribution of ZIP3-level clustering. All analyses were conducted using R version 4.5.0 (R Core Team, 2025). LLMs were used to assist analytical code generation; all code and output was reviewed by the authors, who are accountable for the findings.

### Sensitivity Analyses

We conducted several sensitivity analyses to assess robustness of our primary model: (1) exclusion of covariates with higher levels of missingness, and (2) exclusion of comorbidity indicators. Direction and magnitude of key associations were compared with the primary model.

## Results

### Sample Characteristics

Of the 401,450 women in our cohort, 280,595 (69.9%) achieved an MPR of at least 80% during the first year of adjuvant endocrine therapy.

The cohort included patients from all US census regions, age groups, and racial and ethnic categories (Table 2). Compared with national distributions (Appendix Table 1), patients residing in the Northeast and South were overrepresented (24% and 11%, respectively), whereas those in the West were underrepresented (29%).

Non-Hispanic White patients were overrepresented by 9% relative to national incidence estimates, while Hispanic, Black, and Asian or Pacific Islander patients were underrepresented by 23%, 12%, and 43%, respectively.

Age distributions were broadly consistent with expected incidence patterns, with relative underrepresentation of patients younger than 45 years and 85 years or older (41% and 81% less than expected, respectively), and overrepresentation of those aged 65 to 74 years (approx. 15% more than expected).

**Table 2.** Sample Characteristics, by adherence outcome.

| Characteristic |  | Overall<br>N = 401,450 | Non- adherent<br>N = 120,855 | Adherent<br>N = 280,595 |
| --- | --- | --- | --- | --- |
| **Census region** (p < 0.001)* |  |  |  |  |
|  | Midwest | 94,111 | 27,183 (29%) | 66,928 (71%) |
|  | Northeast | 90,797 | 25,069 (28%) | 65,728 (72%) |
|  | South | 152,402 | 47,434 (31%) | 104,968 (69%) |
|  | West | 64,140 | 21,169 (33%) | 42,975 (67%) |
| **Patient Race/Ethnicity** (p < 0.001)* |  |  |  |  |
|  | Asian or Pacific Islander | 10,328 | 2,869 (28%) | 7,459 (72%) |
|  | Black or African American | 35,238 | 11,145 (32%) | 24,093 (68%) |
|  | Hispanic or Latino | 29,542 | 8,930 (30%) | 20,612 (70%) |
|  | Missing / Unknown | 53,166 | 16,452 (31%) | 36,624 (69%) |
|  | Other | 13,868 | 4,309 (31%) | 9,559 (69%) |
|  | White | 259,308 | 77,060 (30%) | 182,248 (70%) |
| **Insurance Status** (p < 0.001)* |  |  |  |  |
|  | Commercial | 192,112 | 59,555 (31%) | 132,557 (69%) |
|  | Medicare | 209,338 | 60,797 (29%) | 148,541 (71%) |
| **Age group** (p < 0.001)* |  |  |  |  |
|  | Less than 50 | 40,406 | 15,794 (39%) | 24,612 (61%) |
|  | 50-59 | 92,069 | 28,538 (31%) | 63,531 (69%) |
|  | 60-69 | 135,262 | 37,691 (28%) | 97,571 (72%) |
|  | 70-79 | 103,646 | 29,083 (28%) | 74,563 (72%) |
|  | 80+ | 30,067 | 9,749 (32%) | 20,318 (68%) |

\* Pearson's Chi-squared test

### Model Performance

The mixed-effects model demonstrated modest discrimination, with an ROC-AUC of 0.59 in the held-out test set. ZIP3-level clustering contributed minimally to explained variance (intraclass correlation approximately 0.2%), indicating limited residual geographic heterogeneity after adjustment for observed covariates.

### Sociodemographic and Clinical Factors Associated with Adherence

Consistent with other studies, age was strongly associated with adherence (Figure 1). Compared with patients younger than 50 years, those aged 50 to 59 years (aOR, 1.24; 95% CI, 1.19-1.29), 60 to 69 years (aOR, 1.30; 95% CI, 1.22-1.38), and 70 to 79 years (aOR, 1.17; 95% CI, 1.08-1.28) had higher odds of achieving adherence, whereas patients aged 80 years or older had lower odds (aOR, 0.87; 95% CI, 0.78-0.97). Other studies have found similar patterns [6, 29].

Relative to non-Hispanic White patients, Asian or Pacific Islander patients (aOR, 1.11; 95% CI, 1.05-1.17) and Hispanic or Latino patients (aOR, 1.06; 95% CI, 1.02-1.09) were more likely to adhere, while Black or African American patients were less likely to do so (aOR, 0.95; 95% CI, 0.92-0.98). Patients with commercial insurance had higher odds of adherence compared with those insured through Medicare (aOR, 1.07; 95% CI, 1.04-1.11).

Several preexisting comorbidities were negatively, though modestly, associated with adherence.

**Figure 1.**
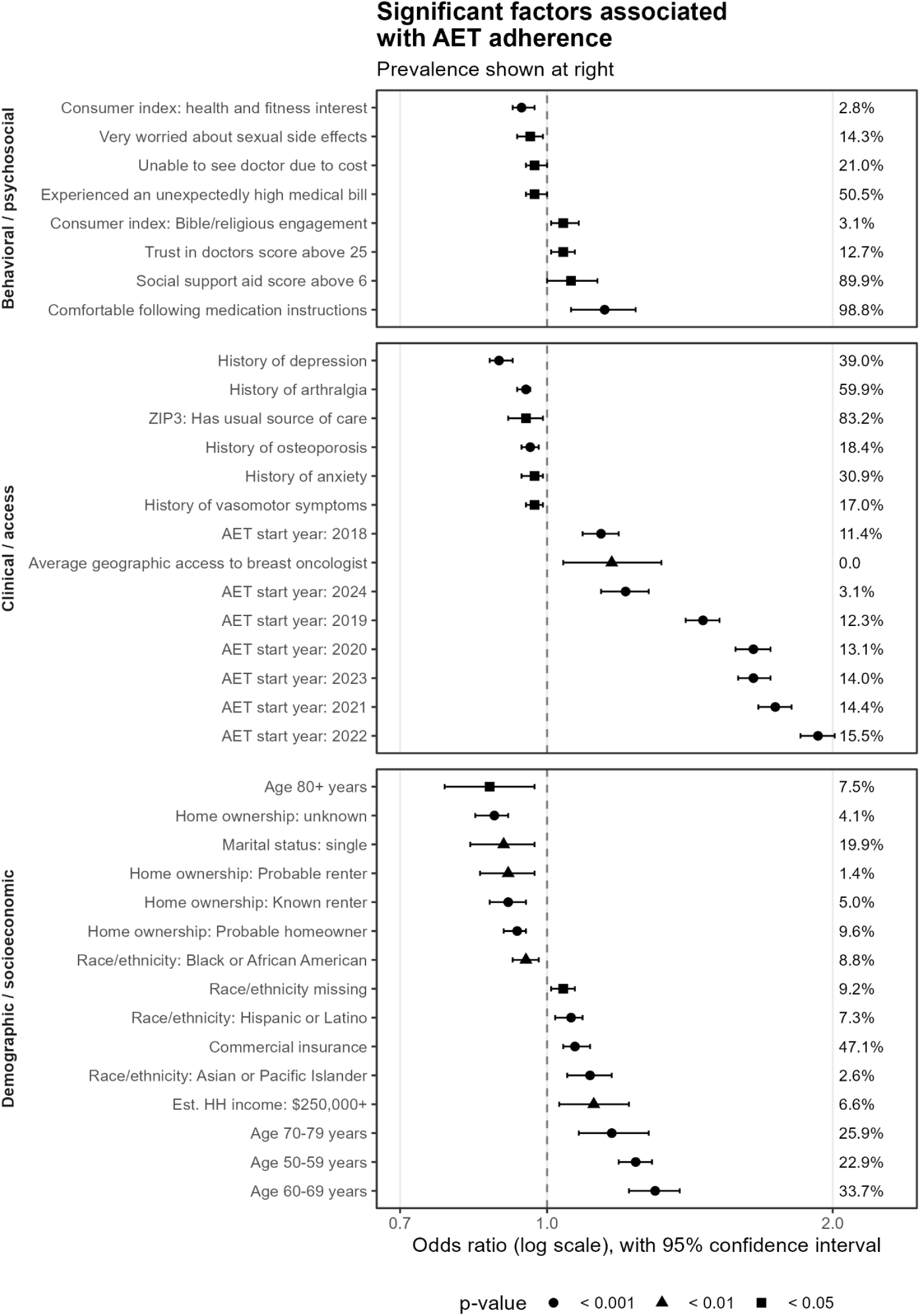
Significant factors associated with adjuvant endocrine therapy (AET) adherence. Adjusted odds ratios (aORs) with 95% confidence intervals are shown for statistically significant predictors from a mixed-effects logistic regression model (n = 401,450). Behavioral, sociodemographic, and clinical predictors are displayed. The reference line at aOR = 1.0 indicates no association. Values greater than 1.0 indicate higher odds of adherence (MPR ≥80%); values less than 1.0 indicate lower odds. Variable mean is expressed as a percentage on the right-hand side. Geographic access to breast oncologists, a continuous variable, was z-scored and therefore has a mean of 0.

### Behavioral Factors Associated with Adherence

After adjusting for clinical and demographic factors, we observed eight significant associations between individual behavioral factors and adherence. These demonstrated clinically meaningful associations, though the magnitude of association was generally modest. Greater concern about sexual side effects was associated with lower adherence (aOR, 0.96; 95% CI, 0.93-0.99). Cost-related access barriers were also associated with reduced adherence, including having been unable to see a doctor because of cost (aOR, 0.97; 95% CI, 0.95-1.00) and experiencing an unexpected high medical bill in the prior two years (aOR, 0.97; 95% CI, 0.95-1.00).

In contrast, several behavioral factors were associated with higher adherence, even after adjusting for covariates such as education, homeownership, and income. Patients reporting comfort with following medication instructions had substantially higher odds of adherence (aOR, 1.15; 95% CI, 1.06-1.24). Higher levels of instrumental social support, as measured by the aid subscale from the Norbeck Revised Social Support Questionnaire [30], were also positively associated with adherence (aOR, 1.06; 95% CI, 1.00-1.13). The two other subscales of the Questionnaire - Affect and Affirm - did not emerge as significant predictors. Greater geographic access to breast oncologists, proxied as the average distance to the 5 nearest providers, was associated with increased adherence (aOR, 1.17; 95% CI, 1.04-1.32).

Consumer interest in Bible-related materials, a proxy for Christian religiosity, was associated with higher adherence (aOR, 1.04; 95% CI, 1.01-1.08). Unexpectedly, higher interest in health and fitness was associated with lower adherence (aOR, 0.94; 95% CI, 0.92-0.97). That this association persisted after adjustment for race, age, and other covariates suggests a complex relationship between health orientation and persistence with long-term adjuvant therapy.

### Common behavioral profiles of adherence drivers

While individual associations were modest, the cumulative impact of behavioral indicators was substantial. The eight behavioral predictors formed several recurrent profiles, with the 10 most common profiles accounting for over 70% of the population. The two most prevalent profiles alone represented over 40% of patients (Figure 2). Relative odds of adherence ranged from 0.92 (95% CI, 0.89–0.95) to 1.04 (95% CI, 1.01–1.07).

**Figure 2:**
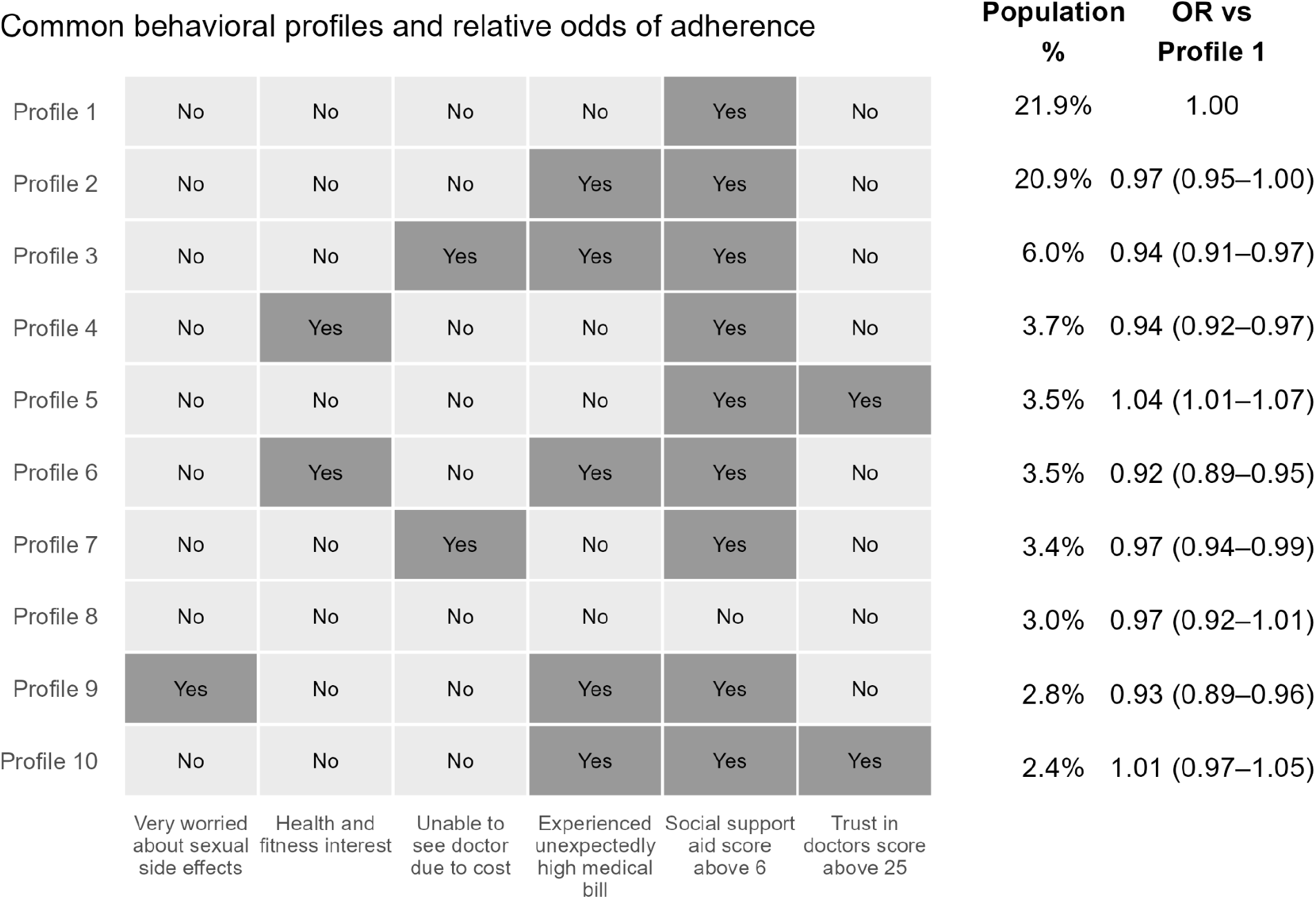
Common behavioral profiles and relative odds of adherence. Rows represent profiles ordered by their prevalence in the patient population. Columns represent behavioral factors identified as significantly associated with adherence. Shading indicates the presence or absence of each factor in a segment. The odds ratio (OR) reflects the odds of adherence compared to the most common profile, with 95% confidence intervals simulated via Monte Carlo draws from the multivariate normal distributions of the behavioral estimates from the fully specified model. Not shown are two features that had a significant association with adherence but were all “Yes” (“Comfortable with following medication instructions”) or all “No” (“Religiosity”) in the ten most prevalent behavioral profiles.

The most common profile - reflecting 22% of the patient population - was characterized by the absence of cost barriers, no concern about sexual side effects, high social support, and average levels of trust in clinicians. The other profiles showed modest but consistent differences in adherence propensity. Profiles characterized by cost-related barriers showed consistently lower adherence propensity, particularly when both types of cost barriers were present. Concern about sexual side effects was also associated with lower adherence. In contrast, higher trust in clinicians was associated with modestly higher adherence in the absence of other barriers. Differences related to social support and fitness orientation were smaller in magnitude and less consistently distinguishable.

### Sensitivity Analyses

Across both sensitivity analyses, the direction and relative magnitude of key associations were preserved, with no sign reversals or material deviations from the primary model estimates. These findings support the robustness of the observed associations to alternative variable specifications and covariate sets.

## Discussion

In this large national cohort, 30.1% of women receiving adjuvant endocrine therapy did not meet recommended adherence criteria during the first year of treatment. By combining large-scale claims data with individual-level behavioral data, we found that cost-related barriers, health literacy proxies, social support, religiosity, fitness intent, and geographic access to breast oncology care were independently associated with adherence after adjusting for a broad set of demographic and clinical factors. We identified patient profiles defined by these behavioral factors, which showed that similar adherence risk can arise from substantially different behavioral drivers.

In real-world settings, patients possess a constellation of preferences and behavioral risks that affects their adherence [11, 12]. Past work has mostly focused on isolating individual risk factors [11, 12]. We were able to extend this work by analyzing a wide set of behavioral factors simultaneously, adjusting for clinical and sociodemographic factors, in a national sample of over 400,000 patients. Doing so, we found that individual associations were modest and generally in line with prior work. Cost-related care disruptions were associated with lower adherence, consistent with evidence that perceived affordability and anticipated quality-of-life tradeoffs operate independently of insurance coverage [16, 17]. Aid-related social support was associated with higher adherence, while emotional support did not emerge as a significant correlate; this is consistent with prior findings that tangible rather than affective support has more measurable effects on treatment behavior [13, 31]. Concern about sexual side effects and psychological symptom burden were associated with lower adherence, extending prior work on anxiety and AET persistence [32]. An unexpected finding was an inverse association between health-and-fitness orientation and adherence. This suggests that a general health-conscious identity does not reliably translate into persistence with long-term endocrine therapy, a pattern consistent with evidence that exercise-focused health behaviors and pharmacological adherence may represent distinct motivational pathways [11, 33, 34].

Our analysis of common behavioral profiles found distinct, clinically interpretable patterns that translate into predictable net associations with adherence and collectively account for over 70% of patients. Compared to the predominant profile, the remaining nine showed relative odds of adherence ranging from 0.92 to 1.04. Though modest at individual levels, the effect is meaningful at system or population scale. The critical observation is that profiles with entirely different pathways can have similar associations with adherence in the same way that multiple clinical risk factors contribute to overall disease risk. For example, a profile featuring two cost-related concerns displays approximately the same association with adherence as one without cost concerns but characterized by a health-and-fitness orientation. This finding may explain, in part, the inconsistent results of previous adherence-targeted interventions [8, 35, 36], even among demographically and clinically similar cases. Similar aggregate outcomes, different underlying factors.

That compositional heterogeneity has direct implications for intervention design. Programs targeting a single domain, such as financial assistance, information, or access, may benefit patients whose non-adherence stems from that source while leaving others unaffected. Approaches that begin by characterizing patients’ behavioral context, through structured screening, care navigator assessment, or data-informed risk stratification, could better match support to the actual distribution of barriers a given population presents rather than an assumed one. Existing evidence supports the potential value of this orientation: health literacy interventions and patient navigation programs have each shown benefit in specific contexts, though their effects on AET adherence specifically remain insufficiently studied [37, 38].

Several limitations warrant consideration. First, the study is observational and cannot establish causal relationships between behavioral factors and adherence. Second, behavioral propensity scores represent modeled probabilities rather than direct measurements of individual beliefs; measurement error likely attenuates associations toward the null. Third, the cohort was limited to insured patients with observable pharmacy claims, limiting generalizability to uninsured or self-pay populations. Fourth, race and ethnicity were incompletely captured in claims data; although sensitivity analyses suggested robustness to alternative specifications, residual bias is possible. Fifth, our cohort excluded mortality in the first postsurgical year; therefore the cohort has survivorship bias and our estimates of adherence may be biased upwards. Sixth, our study cannot determine whether non-adherence is intentional or unintentional. Finally, adherence was operationalized using a binary MPR threshold, which obscures clinically relevant variation in persistence and timing of discontinuation. This limitation likely contributed to the modest discriminative performance of the model and should be considered when interpreting effect sizes. Despite these limitations, inherent to our approach, the findings extend our understanding of personalized medicine in novel ways.

No single behavioral characteristic fully accounts for adherence behavior. Patients arrive at non-adherence through distinct combinations of cost concerns, social resources, health beliefs, and access barriers, each configuration influencing adherence likelihood in its own way. Recognizing that structure, rather than averaging across it, may be the more productive basis for designing adherence support that works across the complex patient population it must serve.

## Acknowledgements

No acknowledgements.

