## Appendix for "Behavioral profiles associated with adherence to adjuvant endocrine therapy in breast cancer: a retrospective population-based cohort study"

### Appendix Table 1. Cohort representativeness relative to national breast cancer incidence estimates, by age group, race/ethnicity, and census region.

*For each characteristic, expected shares were derived from published breast cancer incidence data. For age and race/ethnicity, expected case counts are based on HR+/HER2− breast cancer cases from the SEER 21 registries (November 2024 submission). For geography, expected shares are based on average annual counts of all invasive breast cancer cases across all US states, reported by state cancer registries via State Cancer Profiles (SEER November 2024 and NPCR 2024 submissions, data years 2018–2022), aggregated to the four US Census Bureau regions; HR+/HER2− stratification is not available at this level. The "ratio observed/expected" column reflects the degree of over- or underrepresentation in the cohort: values above 100% indicate overrepresentation and values below 100% indicate underrepresentation.*

a. Age

| | **Number of SEER 21 cases** | **Expected share** | **Observed share** | **Ratio observed/ expected** |
|---|---|---|---|---|
| 40 to 44 years | 22,173 | 5.4% | 4.0% | 75% |
| 45 to 49 years | 33,657 | 8.2% | 6.0% | 74% |
| 50 to 54 years | 39,256 | 9.5% | 10.0% | 105% |
| 55 to 59 years | 44,941 | 10.9% | 12.9% | 118% |
| 60 to 64 years | 58,289 | 14.2% | 16.7% | 118% |
| 65 to 69 years | 65,207 | 15.8% | 17.0% | 107% |
| 70 to 74 years | 60,687 | 14.7% | 15.4% | 105% |
| 75 to 79 years | 42,700 | 10.4% | 10.4% | 100% |
| 80 to 84 years | 25,650 | 6.2% | 6.2% | 100% |
| 85 years and over | 19,344 | 4.7% | 1.2% | 26% |

b. Race/Ethnicity

| | **Number of SEER 21 cases** | **Expected share** | **Observed share** | **Ratio observed/ expected** |
|---|---|---|---|---|
| Asian or Pacific Islander | 35,835 | 8.5% | 3.1% | 37% |

| | | | | |
|---|---|---|---|---|
| Black or African American | 42,373 | 10.0% | 10.5% | 105% |
| Hispanic or Latino | 62,007 | 14.7% | 8.8% | 60% |
| White non-Hispanic | 282,590 | 66.8% | 77.5% | 116% |

### c. Geography

| | **Avg. annual cases (SEER+NPCR, 2018–2022)** | **Expected share** | **Observed share** | **Ratio observed/ expected** |
|---|---|---|---|---|
| Northeast | 51,033 | 18.9% | 22.6% | 120% |
| Midwest | 57,850 | 21.4% | 23.4% | 109% |
| South | 102,525 | 37.9% | 38.0% | 100% |
| West | 58,835 | 21.8% | 16.0% | 73% |

### Appendix Table 2. Unadjusted and adjusted odds ratios for behavioral predictors

*Unadjusted (univariate logistic regression) and adjusted (mixed-effects logistic regression with ZIP3 random intercept) odds ratios for behavioral predictors of one-year AET adherence (MPR ≥ 80%; n = 401,450). Predictors are listed alphabetically by label. Continuous scales (e.g., Champion scale, fatalism, social support, trust in doctors) were dichotomized at prespecified cutoff points; "score above X" indicates the upper category. Sociodemographic, clinical, and geographic covariates are not duplicated here.*

| Predictor | Crude OR (95% CI) | Crude p | Adjusted OR (95% CI) | Adjusted p |
|---|---|---|---|---|
| Agrees health is a matter of luck | 0.82 (0.79–0.84) | <0.001 | 0.99 (0.93–1.05) | 0.7008 |
| Agrees health means everything to them | 1.07 (1.05–1.09) | <0.001 | 1.00 (0.97–1.03) | 0.9368 |
| Agrees safety comes first for health | 1.12 (1.10–1.14) | <0.001 | 1.02 (0.99–1.04) | 0.2462 |
| Agrees they have high social support | 1.20 (1.18–1.22) | <0.001 | 0.99 (0.96–1.02) | 0.5886 |
| Agrees they organize life around future health | 1.10 (1.08–1.12) | <0.001 | 1.02 (0.99–1.05) | 0.1415 |
| Agrees they take good care of their body | 1.14 (1.13–1.16) | <0.001 | 1.01 (0.99–1.04) | 0.3612 |
| Breast cancer knowledge score: 4 correct | 1.13 (1.11–1.15) | <0.001 | 1.00 (0.98–1.03) | 0.7717 |
| Champion barriers subscale score above 15 | 0.86 (0.85–0.88) | <0.001 | 1.00 (0.98–1.03) | 0.7897 |
| Champion scale total score above 50 | 0.81 (0.79–0.82) | <0.001 | 0.99 (0.96–1.02) | 0.5925 |
| Champion susceptibility subscale score above 6 | 1.11 (1.09–1.13) | <0.001 | 0.98 (0.95–1.01) | 0.1317 |
| Comfortable completing medical forms | 1.13 (1.09–1.17) | <0.001 | 0.98 (0.94–1.03) | 0.4565 |
| Comfortable finding health information from multiple sources | 1.20 (1.17–1.24) | <0.001 | 1.00 (0.96–1.05) | 0.9048 |
| Comfortable following medication instructions | 1.45 (1.37–1.54) | <0.001 | 1.15 (1.06–1.24) | <0.001 |
| Consumer index: Bible/religious engagement | 1.06 (1.04–1.09) | <0.001 | 1.04 (1.01–1.08) | 0.0107 |
| Consumer index: health and fitness interest | 0.99 (0.97–1.01) | 0.1688 | 0.94 (0.92–0.97) | <0.001 |
| Correctly knows overweight increases breast cancer risk | 1.10 (1.09–1.12) | <0.001 | 1.01 (0.99–1.04) | 0.3378 |
| Could not fill prescription due to cost | 0.83 (0.81–0.84) | <0.001 | 0.98 (0.95–1.01) | 0.1354 |
| Experienced an unexpectedly high medical bill | 0.99 (0.98–1.00) | 0.1279 | 0.97 (0.95–1.00) | 0.0227 |
| Fatalism score above 15 | 0.91 (0.89–0.92) | <0.001 | 1.02 (0.99–1.05) | 0.1140 |
| Health perceptions score above 35 | 0.96 (0.94–0.99) | 0.0029 | 1.01 (0.98–1.05) | 0.4103 |

| Predictor | Crude OR (95% CI) | Crude p | Adjusted OR (95% CI) | Adjusted p |
|---|---|---|---|---|
| History of breast cancer in a non-family member | 1.07 (1.05–1.09) | <0.001 | 0.99 (0.96–1.02) | 0.3539 |
| Managing addiction issues is very challenging | 0.71 (0.68–0.74) | <0.001 | 1.00 (0.94–1.06) | 0.9437 |
| Managing general health is challenging | 0.80 (0.78–0.81) | <0.001 | 0.99 (0.96–1.02) | 0.52462 |
| Primary support person: family member or relative | 0.84 (0.83–0.85) | <0.001 | 1.01 (0.95–1.08) | 0.7455 |
| Primary support person: friend | 0.81 (0.79–0.83) | <0.001 | 1.02 (0.95–1.10) | 0.5420 |
| Primary support person: health care provider | 0.83 (0.81–0.86) | <0.001 | 1.03 (0.97–1.09) | 0.4044 |
| Primary support person: spouse or partner | 1.07 (1.06–1.09) | <0.001 | 1.00 (0.97–1.02) | 0.7034 |
| Social support affirmation score above 6 | 1.23 (1.20–1.27) | <0.001 | 0.95 (0.91–1.00) | 0.0532 |
| Social support aid score above 6 | 1.21 (1.19–1.24) | <0.001 | 1.06 (1.00–1.13) | 0.0441 |
| Strongly agrees cancer is meant to be | 1.06 (1.02–1.11) | 0.0073 | 1.02 (0.96–1.08) | 0.5248 |
| Strongly agrees cancer makes them anxious | 0.76 (0.74–0.78) | <0.001 | 0.96 (0.93–1.01) | 0.0871 |
| Strongly agrees cancer makes them feel edgy | 0.92 (0.88–0.96) | <0.001 | 0.99 (0.94–1.04) | 0.6871 |
| Strongly agrees cancer makes them feel scared | 0.86 (0.84–0.89) | <0.001 | 0.98 (0.94–1.03) | 0.4481 |
| Strongly agrees cancer makes them feel uneasy | 0.91 (0.89–0.93) | <0.001 | 1.00 (0.96–1.03) | 0.9443 |
| Strongly agrees cancer prevention is not possible | 0.95 (0.90–0.99) | 0.0217 | 1.06 (0.99–1.13) | 0.0759 |
| Strongly agrees death is unavoidable with cancer | 0.89 (0.85–0.93) | <0.001 | 0.98 (0.92–1.04) | 0.4293 |
| Strongly agrees doctors will make the right diagnosis | 1.02 (1.01–1.05) | 0.0138 | 1.00 (0.98–1.03) | 0.7712 |
| Strongly agrees life pushes them around | 0.76 (0.72–0.79) | <0.001 | 0.98 (0.93–1.04) | 0.5585 |
| Strongly agrees others say they take health risks | 0.78 (0.74–0.82) | <0.001 | 0.98 (0.90–1.06) | 0.5500 |
| Strongly agrees patients are not victims of rising health care costs | 0.97 (0.94–0.99) | 0.0163 | 0.98 (0.94–1.03) | 0.4522 |
| Strongly agrees patients receive sufficient information | 1.05 (1.03–1.08) | <0.001 | 0.99 (0.96–1.03) | 0.6503 |
| Strongly agrees they have taken risks with their health | 0.78 (0.75–0.81) | <0.001 | 1.01 (0.96–1.06) | 0.6697 |

| Predictor | Crude OR (95% CI) | Crude p | Adjusted OR (95% CI) | Adjusted p |
|---|---|---|---|---|
| Strongly agrees they would give things up for health | 0.95 (0.93–0.97) | <0.001 | 0.99 (0.96–1.03) | 0.6372 |
| Strongly agrees things tend to go wrong for them | 0.73 (0.70–0.77) | <0.001 | 1.00 (0.93–1.07) | 0.9859 |
| Total social support score above 24 | 1.05 (1.04–1.07) | <0.001 | 0.99 (0.97–1.01) | 0.2639 |
| Trust in doctors score above 25 | 1.09 (1.06–1.11) | <0.001 | 1.04 (1.01–1.07) | 0.0227 |
| Two or more self-management domains rated very challenging | 0.80 (0.79–0.82) | <0.001 | 1.00 (0.97–1.04) | 0.8618 |
| Unable to see doctor due to cost | 0.85 (0.83–0.86) | <0.001 | 0.97 (0.95–1.00) | 0.0452 |
| Very high immediate-help social support | 1.18 (1.16–1.19) | <0.001 | 1.03 (1.00–1.07) | 0.0675 |
| Very worried about fatigue as a side effect | 0.89 (0.88–0.91) | <0.001 | 1.01 (0.99–1.04) | 0.3333 |
| Very worried about hot flashes/sweating as a side effect | 0.81 (0.79–0.82) | <0.001 | 0.97 (0.94–1.01) | 0.1278 |
| Very worried about sexual side effects | 0.79 (0.78–0.81) | <0.001 | 0.96 (0.93–0.99) | 0.0145 |

### Appendix Table 3: Sensitivity Analysis 1 - Excluding covariates with high missingness

*Primary model against a reduced model (columns 4, 5) dropping number of children in the household, race/ethnicity, educational status, and marital status, all of which had elevated missingness rates as reported (Supplementary Table 2).*

| Predictor | aOR (95% CI) primary | p | aOR (95% CI) reduced | p | Significant in | Sign flip |
|---|---|---|---|---|---|---|
| Comfortable following medication instructions | 1.146 (1.061, 1.237) | <0.001 | 1.153 (1.068, 1.244) | <0.001 | both | |
| Instrumental social support (Norbeck aid) | 1.068 (1.005, 1.136) | 0.034 | 1.078 (1.014, 1.146) | 0.016 | both | |
| Trust in doctors (TIMHSS > 25) | 1.038 (1.006, 1.071) | 0.020 | 1.036 (1.004, 1.069) | 0.029 | both | |
| Consumer Bible index (religiosity proxy) | 1.044 (1.010, 1.078) | 0.010 | 1.047 (1.013, 1.081) | 0.006 | both | |
| Consumer health-and-fitness index | 0.944 (0.918, 0.971) | <0.001 | 0.950 (0.925, 0.977) | <0.001 | both | |
| Concern about sexual side effects | 0.966 (0.935, 0.997) | 0.032 | 0.963 (0.933, 0.994) | 0.021 | both | |
| Unable to see a doctor because of cost | 0.974 (0.949, 1.000) | 0.049 | 0.971 (0.946, 0.996) | 0.026 | both | |
| Unexpected high medical bill | 0.972 (0.951, 0.994) | 0.011 | 0.976 (0.955, 0.997) | 0.028 | both | |
| Has a usual source of care | 0.949 (0.910, 0.990) | 0.015 | 0.955 (0.915, 0.996) | 0.033 | both | |
| Geographic access to breast oncologists | 1.167 (1.039, 1.311) | 0.009 | 1.190 (1.057, 1.340) | 0.004 | both | |
| Could not fill prescription (cost) | 0.979 (0.951, 1.007) | 0.143 | 0.971 (0.944, 0.999) | 0.040 | sensitivity only | |
| Immediate help available | 1.029 (0.996, 1.063) | 0.084 | 1.049 (1.016, 1.082) | 0.003 | sensitivity only | |
| Safety comes first for health | 1.017 (0.991, 1.043) | 0.209 | 1.028 (1.002, 1.054) | 0.034 | sensitivity only | |

*All ten significant non-clinical predictors remain significant, in the same direction and with closely similar magnitudes. Three behavioral predictors that were not significant in the primary model reached significance in the reduced specification. Because the point estimate moved with little change to precision, we interpret these as a consequence of removing the sociodemographic covariates rather than as additional findings, and retain the fully adjusted model as primary. This analysis supports the robustness claim as written.*

### Appendix Table 4: Sensitivity Analysis 2 - Excluding comorbidities

*Primary model against the same model with the six pre-index comorbidity indicators removed.*

| Predictor | aOR (95% CI) primary | p | aOR (95% CI) Without comorbidities | p | Change |
|---|---|---|---|---|---|
| Comfortable following medication instructions | 1.146 (1.061–1.237) | <0.001 | 1.139 (1.055–1.230) | <0.001 | -0.6% |
| Instrumental social support (Norbeck aid) | 1.065 (1.002–1.132) | 0.044 | 1.068 (1.005–1.136) | 0.034 | +0.3% |
| Trust in doctors (TIMHSS > 25) | 1.037 (1.005–1.070) | 0.023 | 1.037 (1.004–1.070) | 0.025 | -0.1% |
| Consumer Bible index (religiosity proxy) | 1.043 (1.010–1.078) | 0.011 | 1.045 (1.011–1.079) | 0.009 | +0.1% |
| Consumer health-and-fitness index | 0.943 (0.918–0.970) | <0.001 | 0.936 (0.911–0.963) | <0.001 | -0.8% |
| Concern about sexual side effects | 0.961 (0.931–0.992) | 0.015 | 0.962 (0.932–0.993) | 0.016 | +0.1% |
| Unable to see a doctor because of cost | 0.974 (0.949–0.999) | 0.045 | 0.974 (0.949–1.000) | 0.050 | +0.1% |
| Unexpected high medical bill | 0.975 (0.954–0.996) | 0.023 | 0.976 (0.955–0.997) | 0.028 | +0.1% |
| Has a usual source of care | 0.950 (0.910–0.990) | 0.016 | 0.951 (0.911–0.992) | 0.019 | +0.1% |
| Geographic access to breast oncologists | 1.174 (1.044–1.319) | 0.007 | 1.156 (1.028–1.298) | 0.015 | -1.5% |
| Depression (pre-index) | 0.894 (0.873–0.916) | <0.001 | removed | — | — |
| Arthralgia (pre-index) | 0.947 (0.931–0.964) | <0.001 | removed | — | — |
| Osteoporosis (pre-index) | 0.959 (0.939–0.980) | <0.001 | removed | — | — |
| Anxiety (pre-index) | 0.968 (0.944–0.993) | 0.013 | removed | — | — |
| Vasomotor symptoms (pre-index) | 0.974 (0.953–0.995) | 0.016 | removed | — | — |
| Sexual dysfunction (pre-index) | 0.954 (0.837–1.087) | 0.477 | removed | — | — |

*All ten significant non-clinical predictors remain significant, in the same direction and with closely similar magnitudes. No additional predictors rose to a significant level; this analysis supports the robustness claim as written.*

### Appendix: BehavioralPulse Feature Development

BehavioralPulse is a large-scale social and behavioral dataset developed to estimate latent attitudes, beliefs, preferences, and perceived barriers relevant to healthcare behaviors at the individual level across the United States. In this study, BehavioralPulse was used to generate individual-level behavioral propensity scores specifically related to breast cancer worries, mammography beliefs, side-effect concerns, social support, fatalism, and trust in healthcare. These features were then linked to medical claims data using a privacy-preserving record linkage approach.

#### Primary survey data collection

Breast cancer-specific BehavioralPulse features are derived from a proprietary, nationally representative survey of 6,000 U.S. women of age 40+. The survey was designed using established behavioral science frameworks and oncology expertise to capture constructs that are not observable in administrative or clinical data. These include breast cancer worries, mammography beliefs and barriers, side-effect concerns, social support, fatalism, health literacy, and trust in healthcare.

Surveys were fielded in English and Spanish using high-quality national panels. Survey items were structured to support binary or categorical measurement and downstream predictive modeling. Each item was treated as a distinct behavioral outcome, resulting in 362 candidate breast cancer-relevant variables that served as labeled targets for model training.

#### Predictive modeling and feature generation

To scale survey responses beyond the sampled survey population, we trained a set of supervised machine learning models, one per behavioral construct, using survey responses as ground truth labels. Predictor variables were drawn from a large, individual-level consumer and demographic dataset covering over 260 million U.S. adults, licensed from a third-party vendor. These predictors include both observed attributes (e.g., age, household composition, residential geography), modeled consumer characteristics, and proprietary neighborhood metrics. Race and ethnicity were not used as predictors in the behavioral models.

For each binarized behavioral outcome, we trained models on a subset of survey respondents (70%) and evaluated on held-out test data (30%). Multiple model classes were evaluated depending on outcome prevalence and complexity, with hyperparameters tuned to optimize out-of-sample discrimination and calibration.

#### Model performance and quality assurance

All BehavioralPulse models undergo standardized quality assurance prior to inclusion in analytic workflows. Model discrimination was assessed using the area under the receiver operating characteristic curve (ROC-AUC) on held-out test data. For breast cancer-related features used in this study, ROC-AUC values typically ranged from approximately 0.65 to above 0.90, depending on outcome prevalence and construct complexity.

Calibration was assessed using reliability curves to ensure alignment between predicted probabilities and observed response frequencies across the score distribution. Models exhibiting poor calibration or unstable performance were excluded. For selected constructs with available external benchmarks, we compared aggregated predictions against public datasets at appropriate geographic or population levels to assess plausibility.

#### Population-scale estimation

Validated models were applied to a national consumer dataset to generate behavioral propensity scores for over 45 million U.S. women aged 40+. The final model outputs are individual-level propensity scores ranging from 0 to 1, representing the estimated probability that an individual holds a given belief or concern. This process yielded a dense, individual-level behavioral feature set. These features constitute the BehavioralPulse behavioral layer used in the present analysis. For use in the regression analyses described in the main manuscript, these propensities were binarized at a threshold that results in the known population frequency of the underlying feature to facilitate interpretability of the odds ratios.

BehavioralPulse is designed to reflect population dynamics over time. The underlying consumer dataset is refreshed monthly to capture e.g. residential mobility and household changes, while behavioral models are updated on an as-needed basis.

#### Data linkage and privacy protection

BehavioralPulse features were linked to medical claims data using a privacy-preserving, hash-based tokenization approach [37, 38]. Individual identifiers were converted into encrypted tokens prior to linkage, ensuring that no personally identifiable information was exchanged or exposed. This linkage enabled the integration of behavioral propensities with observed breast cancer diagnoses while maintaining compliance with applicable data protection and privacy standards.

#### Interpretation and limitations

BehavioralPulse features are modeled probabilities rather than direct observations of individual beliefs or behaviors. They should therefore be interpreted as estimates of latent propensities that complement, rather than replace, observed clinical and administrative data. Prediction error in these estimates is expected to attenuate associations toward the null, implying that observed effect sizes likely represent conservative estimates of the influence of behavioral factors.
